# Covid-19 social distancing: when less is more

**DOI:** 10.1101/2021.12.07.21267415

**Authors:** C. Neuwirth, C. Gruber

## Abstract

Covid-19 is the first digitally documented pandemic in history, presenting a unique opportunity to learn how to best deal with similar crises in the future. In this study we have carried out a model-based evaluation of the effectiveness of social distancing, using Austria and Slovenia as examples. Whereas the majority of comparable studies have postulated a negative relationship between the stringency of social distancing (reduction in social contacts) and the scale of the epidemic, our model has suggested a sinusoidal relationship, with tipping points at which the system changes its predominant regime from ‘less social distancing – more cumulative deaths and infections’ to ‘less social distancing – fewer cumulative deaths and infections’. This relationship was found to persist in scenarios with distinct seasonal variation in transmission and limited national intensive care capabilities. In such situations, relaxing social distancing during low transmission seasons (spring and summer) was found to relieve pressure from high transmission seasons (fall and winter) thus reducing the total number of infections and fatalities. Strategies that take into account this relationship could be particularly beneficial in situations where long-term containment is not feasible.

## 1. Introduction

Following the global spread of the new SARS-CoV-2 coronavirus, most governments have decided to impose restrictions on the population (Ebrahim et al., 2020) with the objective of reducing social contacts and preventing epidemic peaks with the potential to overwhelm national health-care systems (Anderson et al., 2020). These restrictions have involved social distancing by, for example, banning large gatherings, closing schools and shops, restricting international travel, and limiting internal mobility. Understanding whether or not the social contact reduction has had the desired effect is critical not only in view of the large societal and economic costs (Flaxman et al., 2020), but also because of the predicted negative impacts on mental health (Giuntella et al., 2021).

Previous studies have shown that such measures have been able to restrict the growth of the epidemic (Flaxman et al., 2020), that mortality rates have been suppressed as a result of early decisions to close schools, public events and state borders (Widimsky et al., 2020), and that social distancing has saved lives (Greenstone & Nigam, 2020; Thunström et al., 2020). These studies have relied on data collected during the early stages of the pandemic, when the stringency and effectiveness of social distancing measures were both high.

Limiting social contacts over the long term may, however, be undermined by the previously-mentioned negative societal and economic effects associated with strictly enforced social contact reduction and isolation. As shown in a previous exploratory study (Neuwirth et al., 2020), if social distancing cannot be sustained over a sufficient length of time (i.e. from outbreak until vaccination and herd immunity), a large second wave of outbreaks can negate the mitigating effects of previously imposed restrictions. This implies that less stringent social distancing may in some instances yield better results than more stringent social distancing in terms of reducing the number of fatalities, since less stringent social distancing can be maintained over longer periods of time.

The same study also revealed that the magnitude of outbreaks that occur following the lifting of restrictions increases according to the stringency of previously applied social distancing, i.e. the suppression of outbreaks through social contact reduction preserves the epidemic potential. Explosive outbreaks following an untimely termination of stringent social distancing are likely to increase case fatality rates if the surge in the number of infected patients exceeds national medical capabilities (Rajgor et al., 2020). In such situations, less stringent social distancing measures may be more effective in curbing the number of fatalities, not only because they can be applied over longer periods of time but also because if they are lifted prematurely the resulting outbreaks are likely to be smaller.

We investigated these hypotheses using a model-based systems analysis. The objectives of this research were (A) to calibrate a mathematical compartment model against one-year long epidemiological time series for Austria and Slovenia, (B) to simulate hypothetical no-social-distancing scenarios in order to investigate the added value achieved by reducing social contacts, and (C) to simulate scenarios with less stringent social distancing (increased number of social contacts) during the early stages of the pandemic (spring 2020) in order to evaluate potential long-term benefits of such a strategy in terms of reductions in infections and fatalities.

The first section below provides a detailed discussion of the method and the data. This is followed by a combined results and discussion section that is structured according to the above-mentioned research objectives.

## 2. Basic model structure

The Covid-19 outbreaks in Slovenia and Austria were modeled using the same compartmental model that was used in a previous exploratory investigation into Covid-19 and social distancing (Neuwirth et al., 2020). In order to address the large number of asymptomatic infections (Gao et al., 2020; Mizumoto et al., 2020), as well as potential increases in case fatality rates due to an excess demand for health facilities (Rajgor et al., 2020; Remuzzi & Remuzzi, 2020), the model extends the standard SIR (susceptible-infected-resistant) model and includes the following compartments: susceptible *S(t)*, infected - infection unknown *I(t)*, infected in isolation *II(t)*, resistant symptomatic *RS*(*t*), resistant asymptomatic *RA(t)*, deaths *D(t)*, deaths caused by denied ICU treatment *DL(t)*.

To calculate the proportions of populations in each compartment we made the simplified assumption that the entire population *N* was initially susceptible. Those susceptible become infected over time by

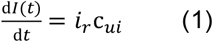

with *i*_*r*_ being the infection rate (i.e. the proportion of contacts between infected and uninfected individuals that result in infections) and *c*_*ui*_ the number of contacts between infected and uninfected, which is calculated as

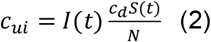

where *c*_*d*_ is the number of social contacts per day.

Asymptomatic infected become resistant as

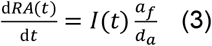

and symptomatic infected are isolated (isolated infected cannot infect others) on confirmation of the disease by

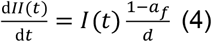

with *a*_*f*_ being the fraction of the infected that are asymptomatic, *d*_*a*_ the duration of asymptomatic infection, and *d* the time between infection and isolation.

Isolated individuals (i.e. home quarantined or hospitalized) die (Eq. 5), die due to a shortage of ICU capabilities (Eq. 6), or become resistant (Eq. 7).

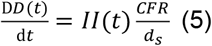

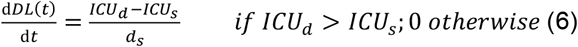

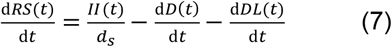

The new parameters in these equations are *d*_*s*_: the duration of distinct symptomatic sickness, *CFR*: the case fatality rate, *ICU*_*d*_: the intensive care demand, and *ICU*_*s*_: the intensive care supply.

To calculate the intensive care demand, we take the critical fraction of *II(t)* that requires admission to intensive care. This fraction is denoted as *c*_*f*_.

We complemented this simple model realization with Google Mobility inputs and recorded ultraviolet light intensities to approximate variations in the number of daily social contacts *c*_*d*_ and variations in seasonal transmissibility. The effects that these inputs had on transmission dynamics were systematically parametrized by means of a Powell optimizer, in order to calibrate the model against statistical records. To test whether alternative calibrations would yield a similar model fit, we conducted 100,000 additional Monte Carlo simulation runs per country. A full description of our approach to consider seasonality and social contacts in the model can be found in the following sections 2.1 and 2.2.

### 2.1 Seasonality

Covid-19 seasonality is particularly evident at higher latitudes, where there is greater seasonal variation in environmental indicators (X. Liu et al., 2021). However, the causal explanation for seasonality remains unclear. For instance, seasonal variations in environmental conditions may change the transmissibility of the virus through the germicidal effects of radiation (Seyer & Sanlidag, 2020), or through changes in human social behavior, or alternatively by affecting the immune response and severity of Covid-19 (Kifer et al., 2021). We implemented seasonal forcing as a function of transmissibility and ignored the possible effects of seasonal indicators on immunization, severity, and mortality.

The variety of environmental predictors with the potential to affect Covid-19 transmissibility presents another challenge to model parametrization. Investigations into Covid-19 seasonality have suggested a significant relationship between ultraviolet (UV) light and rates of spread of Covid-19. Multivariate investigations found that UV light had the strongest correlation with Covid-19 growth (Merow & Urban, 2020) and that UV light was the only statistically significant predictor (Carleton et al., 2021) of those investigated (UV light, temperature, and humidity). Similar results in favor of UV light have been obtained by comparing the effects of ozone with those of UV light (To et al., 2021).

We therefore took into account the effects of UV radiation in the model by modifying the infection rate *i*_*r*_ with

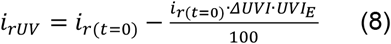

where *i*_*r(t* = 0)_ is the infection rate at model initialization, *ΔUVI* is the change in the daily measured UVI (ultraviolet index) relative to the UVI at model initialization, and *UVI*_*E*_ is the effect of UVI on the infection rate, expressed as a percentage (i.e. per unit increase in UVI, *i*_*r*_ increases by *UVI*_*E*_).

The daily UVI in Equation 8 was obtained from OpenWeather API for seven Austrian provincial capitals and the Slovenian capital Ljubljana. The requested geographic locations correspond to European capitals as provided by EFRAINMAPS. The Austrian local data is arithmetically averaged.

### 2.2 Social contact reduction

The behavior of individuals affects the dynamics of the epidemics, and vice versa. When an outbreak occurs, social contacts are often constrained by governmental regulations but reductions in social contacts can also occur spontaneously as individuals respond to news from public sources about the spread of the disease (Blendon et al., 2004). This behavior change reduces the average number of new infections produced by each infected individual and the severity of the epidemic, which in turn has an effect on subsequent governmental decisions and public social behavior.

To capture these dynamics during the Covid-19 crisis, social contact surveys have been carried out for countries such as Luxemburg (Latsuzbaia et al., 2020) and the US (Feehan & Mahmud, 2021). The use of survey data in a Covid-19 model is, however, constrained by the limited geographic and temporal coverage of the surveys. Mobility data presents an important alternative proxy for social contacts (Nouvellet et al., 2021). However, uncertainty associated with mobility data used as a proxy arises from the possibility of decoupling between mobility volumes and the number of social contacts that are infectious. Precautionary measures such as wearing a mask or maintaining a distance even when encountering individuals, are likely confounding effects (Gatalo et al., 2021). Investigations into the relationship between transmission and mobility have revealed significant correlations during the early phase of the pandemic but also yielded evidence for a decoupling of transmission from mobility following the relaxation of strict control measures (Nouvellet et al., 2021).

In view of these tradeoffs, we decided to use Google Mobility data for reasons of coverage and transferability. These data reflect the movement of people under six categories: “Parks”, “Residential”, “Grocery and Pharmacy Stores”, “Workplaces”, “Retail and Recreation”, and “Transit Stations”. Mobility volumes are represented as positive and negative percentage changes with respect to a 5-week baseline period (January 3 - February 6, 2020). The full technical details can be obtained from Google’s Community Mobility Reports (Google, n. d.).

Of the available mobility classes, “Retail and Recreation” was considered to be the most appropriate for use as a proxy for variations in the number of social contacts on a cross-national scale. This category includes environments such as restaurants, cafes, shopping centers, theme parks, museums, libraries and movie theatres (Google, n. d.), which are particularly affected by cross-national policies of closing non-essential facilities (Redlberger-Fritz et al., 2021; Zadnik et al., 2020). We therefore used the “Retail and Recreation” mobility class *m* to scale the initial number of social contacts per day *c*_*d(t* = 0)_ as

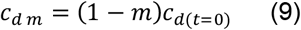

### 2.3 Model parameters

The basic reproduction *R*_0_ is the most fundamental parameter in our model. It represents the number of new infections passed on by an infected person in a completely susceptible population (Dietz, 1993). Seasonality, as well as social distancing in response to an outbreak, have a modifying effect on *R*_0_, i.e. *ΔUVI* ≠ 0 and/or *m* < 0. The resulting modified *R*_0_ is referred to as the effective reproduction *R*, which reduces over time as the pool of susceptible individuals decreases as a result of new infections, i.e.

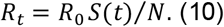

Due to insufficient evidence for a reduced infectiousness of asymptomatic carriers (McEvoy et al., 2021), we assumed equal infectiousness for both symptomatic and asymptomatic carriers and modeled *R*_0_ as

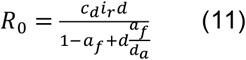

Estimates of the SARS-CoV-2 *R*_0_ vary within a broad range, from 1.4 (Shim et al., 2020) to 8.7 (Linka et al., 2020), which can in part be explained by factors such as differences in social habits, culture (Huynh, 2020; Locatelli et al., 2021), and the methods used to estimate reproductive numbers (Najafimehr et al., 2020). While mathematical models tend to overestimate *R*_0_, the true *R*_0_ for SARS-CoV-2 is expected to be around 2-3 (Y. Liu et al., 2020). This assumption is supported by the results of recent investigations, which suggest a remarkably similar *R*_0_ in most European countries, with an average value of 2.2 (95% CI: 1.9-2.6) (Locatelli et al., 2021). We initialized our model by this number and systematically varied the parameters *c*_*d*(*t*=0)_, *UVI*_*E*_, and *I*_(*t*=0)_ by means of a Powell optimization to fit the modeled cumulative deaths to national statistical records (ECDC, 2020). We then ran Monte Carlo simulations to identify possible alternative model fits in three-dimensional parameter space.

Apart from the *calibration inputs*, we distinguished between *model inputs* and *derivatives* thereof. A complete list of model parameters, together with explanations, is presented in Table 1.

**Table 1:** Model Parameters

| Parameters | Type | Value(s) |
| --- | --- | --- |
| Basic reproduction $R_0$ | Derivative | 2.2 (Locatelli et al., 2021) |
| Infection fatality rate $IFR$ | Derivative | 0.36%<sup>1</sup> |
| Case fatality rate $CFR$ | Model input | 0.53%<sup>2</sup> |
| Initial social contacts per day $c_{d(t=0)}$ | Calibration input | 8<sup>3</sup> |
| Initial infection rate $i_{r(t=0)}$ | Model input | 0.0328<sup>4</sup> |
| Effect of UVI on infection rate $UVI_E$ | Calibration input | 13%<sup>5</sup> |
| Time between infection and isolation $d$ | Model input | 7 days<sup>6</sup> |
| Duration of distinct symptomatic sickness $d_s$ | Model input | 14 days (Xu et al., 2020) |
| Duration of asymptomatic infection $d_a$ | Model input | 14 days |
| Fraction of asymptomatic among infected $a_f$ | Model input | 33% (Oran & Topol, 2021) |
| Outbreak size $I_{(t=0)}$ | Calibration input | AUT 2 SLO 2<sup>7</sup> |
| Number of ICU beds $ICU_s$ | Model input | AUT 2000 SLO 133 (Rhodes et al., 2012) |
| Fraction of confirmed cases that need intensive care $c_f$ | Model input | 1%<sup>8</sup> |

<sup>1</sup> A systematic review by Meyerowitz-Katz & Merone (2020) has indicated that, on average, the IFR is 0.68%. Due to the significant dependence of disease severity on age (Wu et al., 2020), we relied on numbers carried out by Streeck et al. (2020) in a serological study conducted in Germany; a country whose population has a similar median age (45.9) to Austria (43.5) and Slovenia (44.1) (EUROSTAT, n. d.). <sup>2</sup> $CFR = \frac{IFR}{1-a_f}$
<sup>3</sup> This estimate is in line with population surveys conducted in countries such as Germany 7.95, $n=1341$ (Mossong et al., 2008), France 8, $n=2033$ (Béraud et al., 2015), and Luxembourg 7.1, $n=1119$ (Latsuzbaia et al., 2020).
<sup>4</sup> We inserted $i_{r(t=0)} = 0.0328$ into Equation 11 to obtain an $R_0$ value of 2.2.
<sup>5</sup> We used estimates obtained in a Canadian study by To et al. (2021), due to expected similar seasonal dependencies at similar geographic latitudes. A Pakistan study by Adnan et al. (2021) suggested a $UVI_E$ value of 18%.
<sup>6</sup> The selection of this parameter was motivated by a notable similarity between two observations: (a) The duration of presymptomatic infection (6 days) (Xu et al., 2020), and (b) the time between illness onset and reporting (7.1 days on average) (Jung et al., 2020).
<sup>7</sup> The outbreak size corresponds to the number of detected cases at the first emergence of the disease in Austria (Feb. 25<sup>th</sup>) and Slovenia (Mar. 5<sup>th</sup> 2020), according to ECDC (2020).
<sup>8</sup> In Austria, ICU admissions of known infected dropped from a maximum of 7% during the first wave of the pandemic to about 1% from summer 2020 onwards (AGES, n. d.). The high ICU admission rates during the first wave of the pandemic are likely to have been a result of the limited testing capabilities. Austrian records show that the number of tests conducted increased by a factor of more than 20 between April 2020 and February 2021 (ECDC, 2020). We therefore used the lower benchmark for Austria and Slovenia.

## 3. Results and discussion

The calibrated compartment model closely reproduced the trajectories of confirmed daily and cumulative deaths in Slovenia and Austria (see **Figure 1**). Results suggested a higher basic reproduction *R*_0_ for Slovenia than for Austria, due to a larger daily number of social contacts. The *R*_0_ estimates for both countries appear reasonable when compared with results from a cross-European study by Locatelli et al. (2021), which proposed *R*_0_ = 2.21 for Western European countries, and with a previously published comprehensive review by Y. Liu et al. (2020), which indicated that estimates of *R*_0_ in recent studies appear to have stabilized at between 2 and 3. Monte Carlo simulations did not yield any better solution in terms of the root mean square deviation (RMSD) between model outputs and recorded data. The parameters used in Monte Carlo runs with the lowest RMSD converged towards those identified by the Powell optimization (see **Figure 2**). We therefore stuck with the calibration parameters obtained from the Powell optimization.

**Figure 1.**
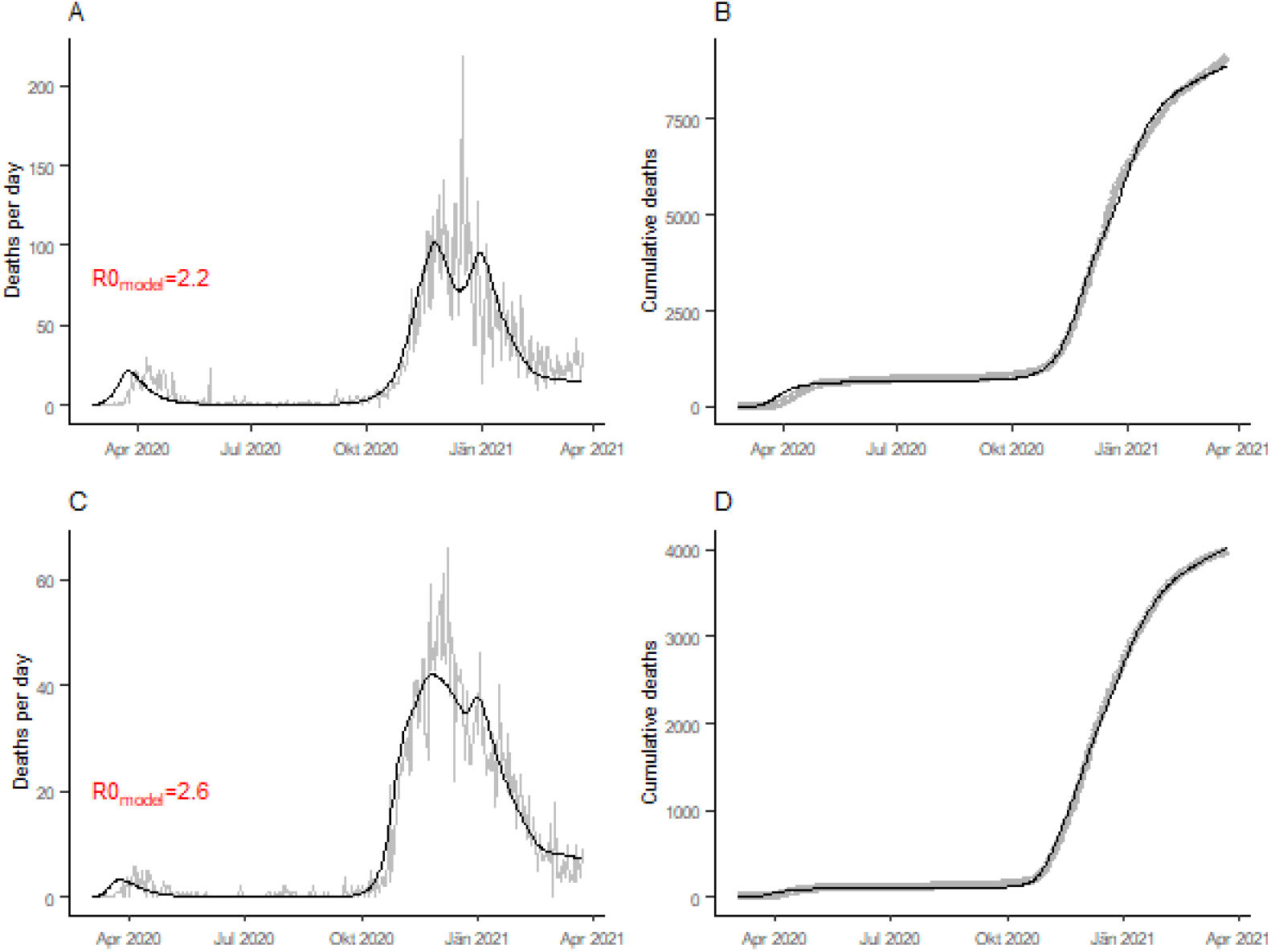
Results of Powell optimization for Austria (A, B) and Slovenia (C, D), showing the calibrated model outputs (black) together with national statistics on Covid-19 deaths (grey). The fitted model parameters are *c*_*d*(*t*=0)_ = 8.28, *UVI*_*E*_ = 8.77 and *I*_(*t*=0)_ = 3829 for Austria with *RMSD =* 121.44, and *c*_*d*(*t*=0)_ = 10.01, *UVI*_*E*_ = 11.9 and *I*_(*t*=0)_ = 1379 for Slovenia with *RMSE =* 30.96. See model parameters in Table 1 for explanation of symbols. The basic reproduction number R0_model_ is estimated from the model’s average effective reproduction *R*_*t*_ between the day of the first detected Covid-19 case and the implementation of national social distancing measures, i.e. between February 25^th^ and March 15^th^ for Austria and between March 5^th^ and March 12^th^ for Slovenia.

**Figure 2.**
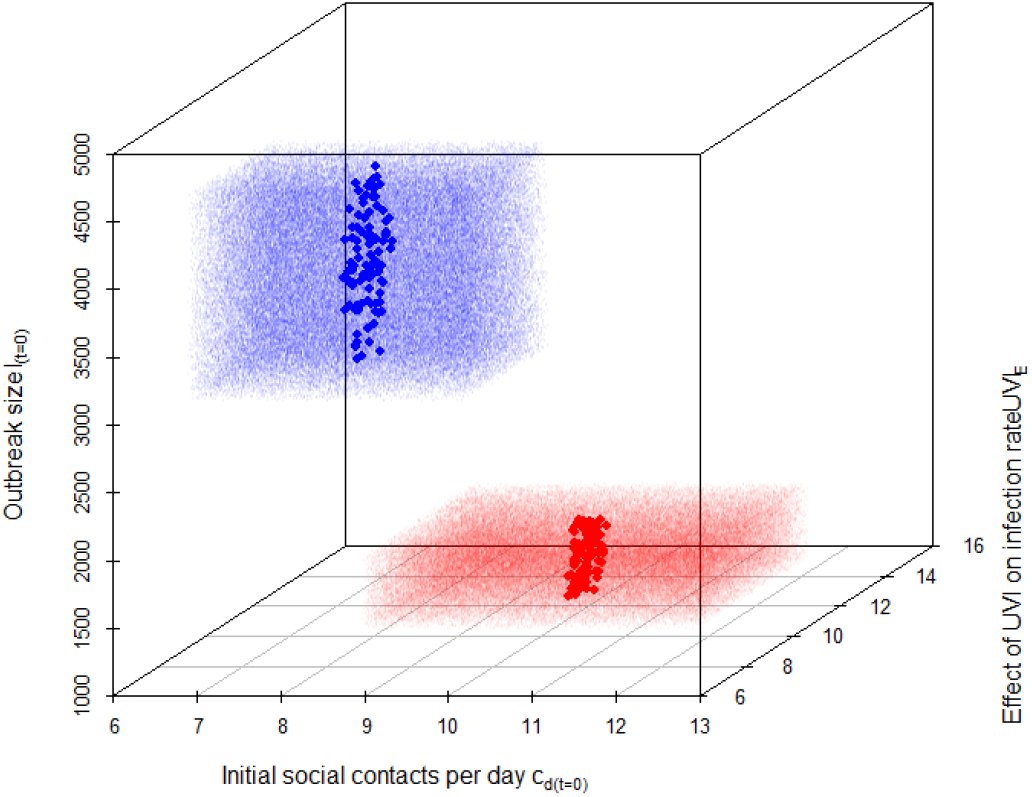
Parameter values from 100,000 (each) Monte Carlo simulation runs: red dots for Slovenia and blue dots for Austria. Random parameters were generated based on a uniform distribution between a minimum value equal to the *′ Fitted model parameter* − 20%′ and a maximum value equal to the *′ Fitted model parameter* + 20%′. Large dots represent the 100 simulation runs with lowest root mean square deviation (RMSD) of modeled from recorded cumulative deaths. Best model fit for Austria: *RMSD*_*min*_ = 122.87 with *c*_*d*(*t*=0)_ = 8.29, *UVI*_*E*_ = 8.80, and *I*_(*t*=0)_ = 3799 and for Slovenia: *RMSD*_*min*_ = 31.23 with *c*_*d*(*t*=0)_ = 10.01, *UVI*_*E*_ = 11.89 and *I*_(*t*=0)_ = 1329. See model parameters in Table 1 for explanation of symbols.

A simulation using these calibration inputs showed the cumulative number of infections by April 23^rd^ 2021 to be 2,148,000 in Austria and 1,203,000 in Slovenia. When compared to the national records of confirmed cases (ECDC, 2020), our model therefore suggests that 75% of infections in Austria and 82% of infections in Slovenia are undocumented. An even lower ascertainment rate of approximately 1 identified case in 12 infections was obtained using a combined data and inference approach for France over a seven-week period from mid-May to the end of June 2020 (Pullano et al., 2021; Shaman, 2020). Other studies (Lau et al., 2021; Li et al., 2020) have estimated similarly high levels of undocumented infections in a variety of countries including France, Italy, Spain, China, and the United States. Due to the gradual extension of national testing capabilities, the proportion of undetected infections to date would now presumably be lower. Nevertheless, our results as well as those in other relevant publications suggest high prevalence and a relatively moderate severity of SARS-CoV-2.

Other studies (Ahmed et al., 2018; Halloran et al., 2008; Kelso et al., 2009) have reported a decline in the effectiveness of social distancing with higher basic reproduction and prevalence. In order to investigate the effectiveness of social distancing in controlling the spread of Covid-19, we compared the fatality numbers from the data-calibrated model run against those from a simulation run without any social distancing. Social contacts in the no-social-distancing scenario were modeled using the Google Mobility baseline data as a proxy for the pre-pandemic situation (see section 2.2).

Results showed that social distancing in Austria and Slovenia greatly mitigated the initial outbreaks (spring outbreaks 2020) but amplified subsequent outbreaks (winter outbreaks 2020/21) (see **Figure 3, A** and C). Although social distancing was shown to be highly effective in reducing fatalities during the early stages of a pandemic, these benefits tend to be lost over the longer term as evident from U-shaped relative fatality curves in **Figure 3, B** and D. These curves represent the ratio of the number of fatalities in scenarios with social distancing to those in scenarios with no social distancing (e.g. a relative fatality value of 0.6 indicates that social distancing reduced fatalities by 40%).

**Figure 3.**
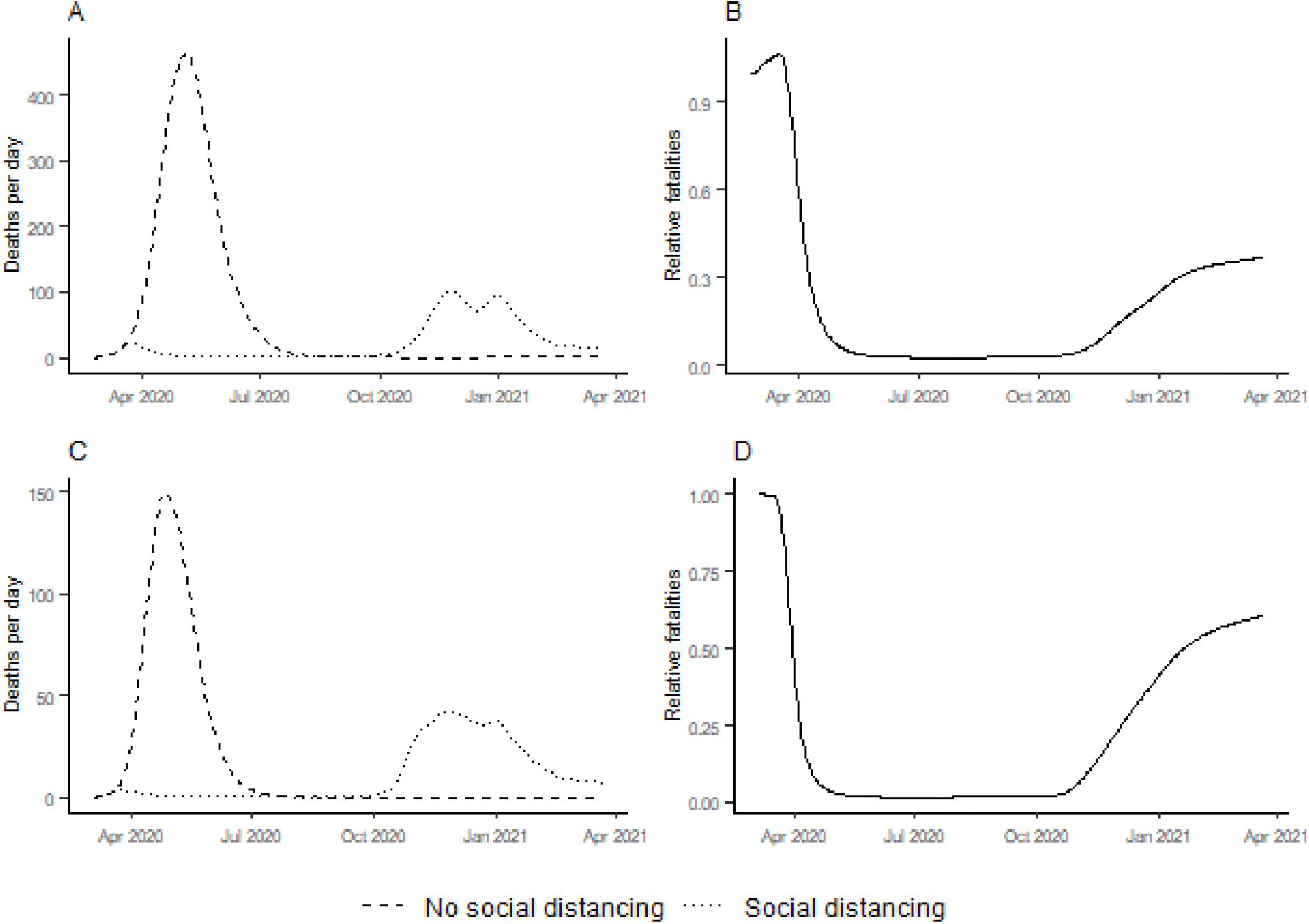
Comparison of simulated outbreaks with and without social distancing in Austria (A, B) and Slovenia (C, D). A and C: simulated outbreaks with social distancing (calibrated against recorded data) and simulated outbreaks with no social distancing. B and D: relative fatalities, i.e. ratio of the number of fatalities in social distancing scenarios to those in no-social-distancing scenarios.

A sensitivity analysis revealed a considerable sensitivity of relative fatality curves to variations in *R*_0_. The application of social distancing in high *R*_0_ scenarios resulted in multiple waves of outbreaks and U-shaped relative fatality curves as depicted in **Figure 3, B** and D (i.e. benefits are lost over the long term), whereas low *R*_0_ scenarios resulted in single outbreaks and L-shaped relative fatality curves. In other words, as anticipated by others, social distancing is effective in low *R*_0_ scenarios but less effective in high *R*_0_ scenarios. Given the projected *R*_0_ values for Slovenia and Austria, social distancing may have reduced fatalities within the study period by about 40% and 63%, respectively.

However, because of uncertainties in the modeling, it is important not to overinterpret these figures. Our results should be viewed as exploratory rather than predictive. Nevertheless, we interpret the observed and modeled patterns as strong indicators that multiple epidemic waves have been caused by the application of social distancing policies under high *R*_0_ conditions. A similar explanation has previously been proposed for the multiple waves of the influenza strain seen in Sydney, Australia, during the 1919 pandemic (Caley et al., 2008).

Moreover, it is speculated that subsequent outbreaks (winter 2020/21) would have been smaller if social distancing during initial outbreaks (spring 2020) was less stringent (i.e. increased number of social contacts in spring 2020). In order to investigate this hypothesis, we simulated less stringent social distancing by gradually increasing recorded mobility volumes (Google Mobility data) by 1% increments in the model for the duration of the initial outbreaks (spring 2020) in Austria and Slovenia (see **Figure 4, A** and D).

**Figure 4.**
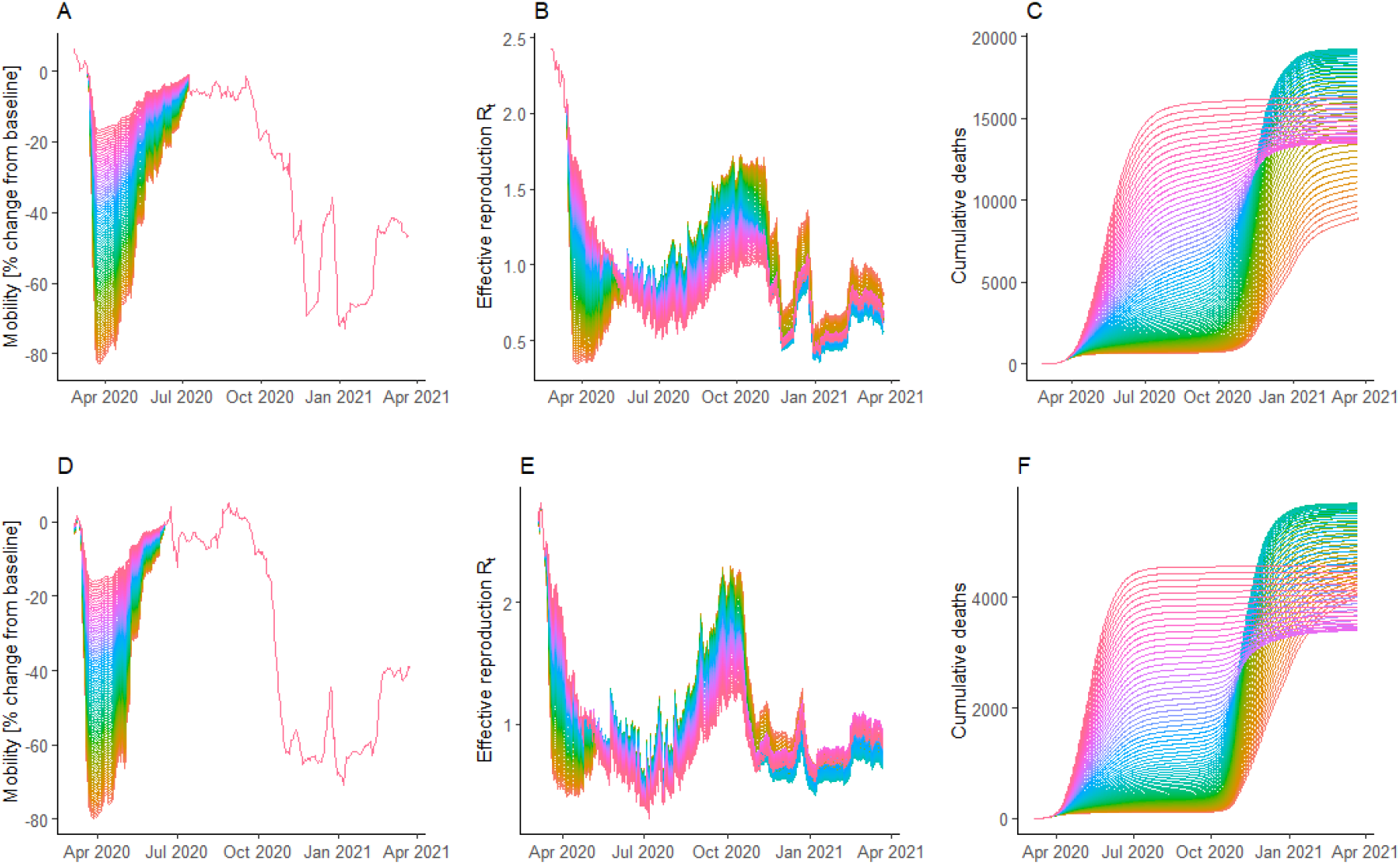
Less stringent social distancing (implemented in the model through a mobility increase) during the initial outbreak and the corresponding effective reproduction and cumulative deaths for Austria (A, B, C) and Slovenia (D, E, F). A and D: Google Mobility volumes, increased in 1% increments (+0%=orange, +80%=pink). B and E: effective reproduction *R*_*t*_ in less stringent (pink) and more stringent (orange) social distancing scenarios. C and F: cumulative deaths in less stringent (pink) and more stringent (orange) social distancing scenarios.

These scenarios revealed the existence of tipping points at which the system changes its dominant regime from ‘less social distancing – more cumulative deaths’ to ‘less social distancing – fewer cumulative deaths’. In order to understand this counterintuitive result, we need to consider the effective reproduction numbers. Effective reproduction *R*_*t*_ is depleted over time as a function of total infections (see Equation 10). As a result, low *R*_*t*_ and small outbreaks follow high *R*_*t*_ and large outbreaks, and vice versa (see **Figure 4, B** and E). Moreover, due to seasonality, *R*_*t*_ was lower in spring 2020 (small outbreaks) than in fall and winter 2020/21 (large outbreaks).

This causes two independent effects that explain why, in some of the scenarios, less stringent social distancing is associated with a smaller cumulative number of deaths (compare **Figure 4A** with **Figure 4 C** as well as **Figure 4 D** with **Figure 4 F**). The first effect (Effect 1) was that less stringent social distancing in spring 2020 led to a balanced allocation of infections among spring 2020 and winter 2020/21 outbreaks (increased number of infections in spring and reduced number of infections in winter), relieving the pressure on national health care systems during the winter of 2020/21 and reducing the overall infection fatality rates (IFR). The second effect (Effect 2) was that less stringent social distancing in spring mitigated high potential winter outbreaks, which overall reduced the total number of infections.

The modeled scenarios showed that these effects are strong enough to reduce the cumulative fatality numbers to below the fatality numbers actually recorded in Slovenia, if modeled social interactions were greatly increased (see **Figure 5**). This was mainly due to a distinctly misbalanced allocation of Slovenian infections, with a small spring outbreak and a much larger winter outbreak, which exceeded national medical capabilities. In model scenarios with less stringent social distancing in spring, this misbalance was corrected and both IFR and fatalities were reduced (Effect 1). Moreover, less stringent social distancing in spring reduced the overall infection numbers in a large range of the simulation runs (Effect 2). A similar effect was previously anticipated in a modeling study by Engelbrecht & Scholes (2021), who predicted large-scale subsequent outbreaks due to initial containment of the disease, the presence of a large pool of susceptible individuals, and favorable conditions in the form of a full winter period.

**Figure 5.**
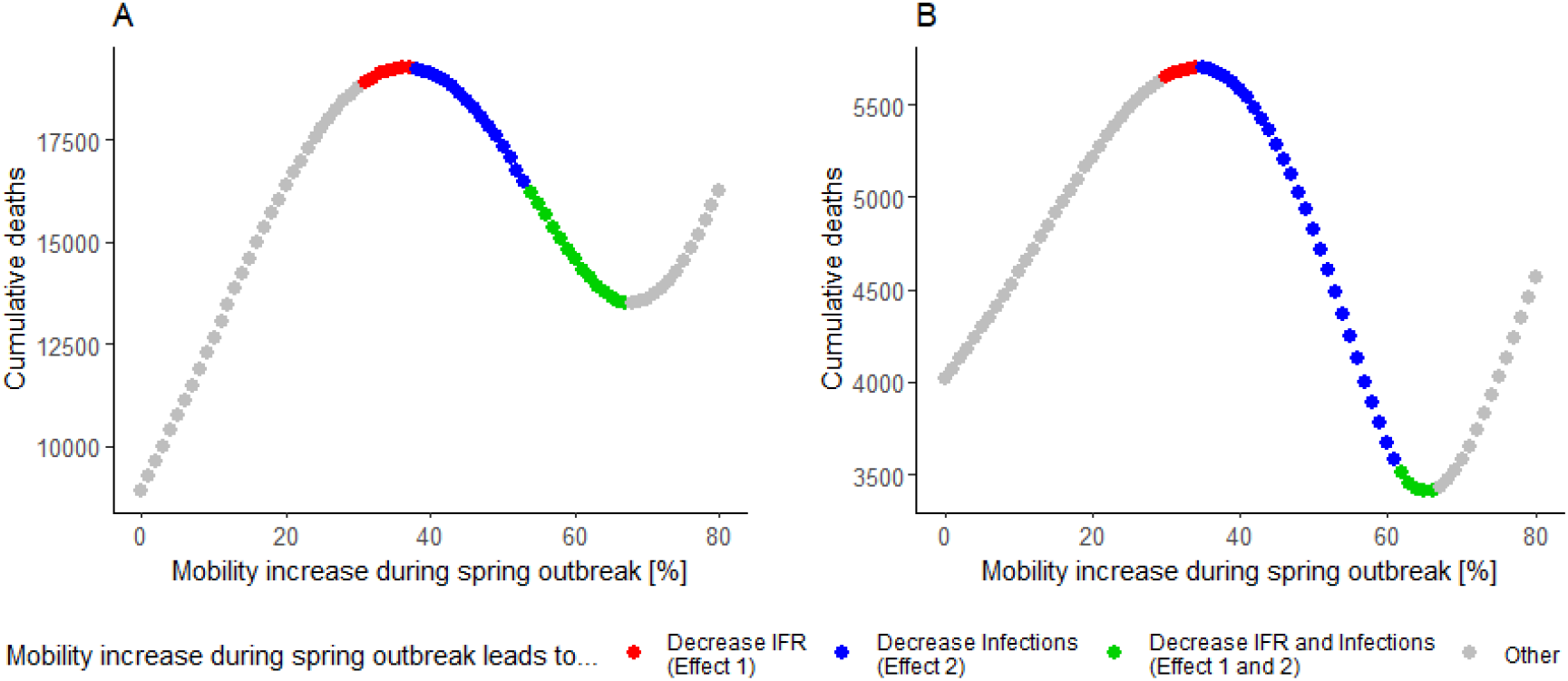
Relationship between stringency of social distancing in spring 2020 (implemented in the model through a mobility increase during the spring outbreaks) and cumulative deaths for Austria (A) and Slovenia (B).

## 4. Conclusion

In this study we applied methods of model-based systems analysis to investigate the effectiveness of social distancing measures in the mitigation of Covid-19, using Austria and Slovenia as examples. Our results showed that contact reduction has drastically curbed infections and fatalities during the early stages of the pandemic. However, these benefits tend to be lost over the long term due to large outbreaks at a later stage of the pandemic, i.e. fatalities in model scenarios with social distancing gradually approximated fatality numbers in scenarios without any social contact reduction. Declining effectiveness of social distancing can be explained by initial containment and the presence of a large pool of susceptible individuals that coincides with elevated transmissibility in fall and winter. A sensitivity analysis showed that an increase in the basic reproduction number *R*_0_ further diminishes the effectiveness of social distancing, which is highly relevant given the presumed gains in transmissibility of newly emerging variant strains of SARS-CoV-2 (Davies et al., 2021).

In view of these preconditions and the expected ineradicable nature of the pathogen, easing social-distancing during low-transmission seasons in order to relieve pressure from high-transmission seasons was found to mitigate large winter outbreaks. This strategy is particularly effective in curbing the overall number of infections and fatalities where health care capabilities are likely being overwhelmed by larger outbreaks, where there is distinct seasonality, and where due to high *R*_0_ long term containment is not feasible.

This effect is of course subject to the condition that reinfection is ignored by the model. A study carried out in the Tyrol (Austria) by Deisenhammer et al. (2021), however, showed a stable and persisting antibody response against SARS-CoV-2 six months after infection suggesting that reinfections are unlikely to be very significant.

The presented analysis suggests reconsidering greedy mitigation strategies that are aimed at minimizing social contacts at all times and that in many cases do not produce an optimal solution. Total eradication and prolonged containment strategies have only proved epidemiologically successful in the long-term for few countries, some of which are characterized by consistently high solar irradiation and negligible seasonality (e.g. Singapore), or by geographic isolation (e.g. New Zealand). In order to further investigate the validity of our hypothesis, we plan to transfer our method to other geographic regions.

## Data Availability

All data produced in the present study are available upon reasonable request to the authors

## Acknowledgments

<empty for review>

## Notes

### Competing Interest Statement

The authors have declared no competing interest.

### Funding Statement

This study did not receive any funding

